# Lessons learnt from implementing FAIRification workflows in diabetes research in Germany

**DOI:** 10.1101/2025.07.01.25330204

**Authors:** Esther Thea Inau, Angela Dedié, Ivona Anastasova, Renate Schick, Brigitte Fröhlich, Michael Roden, Andreas L. Birkenfeld, Martin Hrabě de Angelis, Martin Preusse, Dagmar Waltemath, Atinkut Alamirrew Zeleke

**Affiliations:** Medical Informatics Laboratory, University Medicine Greifswald, Greifswald, Germany; German Center for Diabetes Research (DZD), München-Neuherberg, Germany; German Center for Diabetes Research (DZD), Düsseldorf, Germany; Department of Endocrinology and Diabetology, Medical Faculty and University Hospital Düsseldorf, Heinrich-Heine-University Düsseldorf, Düsseldorf, Germany; Institute for Clinical Diabetology, German Diabetes Center, Leibniz Center for Diabetes Research at Heinrich-Heine-University Düsseldorf, Düsseldorf, Germany; German Center for Diabetes Research (DZD), Tübingen, Germany; Institute for Diabetes Research and Metabolic Diseases of the Helmholtz Zentrum München at the University of Tübingen (IDM), Tübingen, Germany; Department of Diabetology, Endocrinology, and Nephrology, University Clinic Tübingen, Eberhard Karls University Tübingen, Tübingen, Germany; Institute of Experimental Genetics and German Mouse Clinic, Helmholtz Munich, Neuherberg, Germany; Chair of Experimental Genetics, TUM School of Life Sciences (SoLS), Technische Universitäat München, Freising, Germany; Department of Research and Publication Support, University and City Library, University of Cologne, Cologne, Germany

## Abstract

The FAIR principles guide data stewardship towards maximizing the value of scientific data while offering a high level of flexibility to accommodate differences in standards and scientific practices. Research communities have developed and implemented domain-specific workflows to make their data FAIR. This work compares the implementation of a structured generic FAIRification workflow with a domain-specific workflow using the example of metadata captured in diabetes research in Germany and applying the FAIR data maturity model developed by the Research Data Alliance. We show that both workflows require similar resources. Interestingly, the implementation of both workflows led us to achieve the same FAIRness rating. We therefore conclude that the adoptions made in the FAIRification workflow for health research data are useful to improve efficiency but do not necessarily lead to higher FAIRness scores when applied to core data sets. Based on the results of our workflow comparison we identified a list of requirements that should be met for the FAIRification of a core data set regardless of the workflow employed. In the future, FAIR data strategies and infrastructure should be planned and implemented as early as possible in the FAIRification journey. It is anticipated that this comparative analysis will help establish standard operating procedures for the FAIRification of core data sets for health studies.

## Introduction

The FAIR principles guide data stewardship towards improved findability, accessibility, interoperability, and reusability (FAIRness) of research objects [1, 2]. These principles systematically usher research data management (RDM) towards the attainment of maximum value of scientific data and reproducibility of research findings [3]. FAIR RDM strengthens data sharing among heterogeneous data silos and enables collaborative research [4–8].

FAIRification is described as the process of making research objects FAIR [9]. It has been implemented in different sectors of biomedical research from computational biology where the results have helped to improve the quality of modelling pipelines [10] to clinical epidemiology where the results have enabled the combination of metadata on epidemiological studies in Germany [11].

FAIRification typically starts with a FAIR assessment in which a tool is used to determine how FAIR the research object is using a structured assessment [12, 13]. The results of this assessment are used to inform measures that ought to be taken for the research object to become FAIRer. After these measures have been applied, a second assessment is conducted to determine their impact. A higher FAIR score is often the expected outcome of FAIRification [6, 12, 14]. The FAIRification journey, which is often embarked on to fulfil requirements set by funders or research institutions, has been described as intensive [5, 6]. Therefore, work has been done to prescribe simpler steps to improve FAIRness [12, 15]. In this context, FAIRification workflows promise to provide the needed guidance on how to implement the FAIR data principles in a practical and gradual manner [6, 16, 17].

The Research Data Alliance (RDA) has collected maturity indicators and determined evaluation levels that together serve as the assessment criteria for FAIRness [14]. This common set of assessment criteria has been used in FAIR assessments across different scientific communities and has also shown to be useful for other tasks such as the calibration of reporting guidelines for machine learning models in health research [18–21]. The RDA further incorporated these assessment criteria into the FAIR Data Maturity Model (FDMM), which enable comprehensive manual FAIR assessment and guides FAIRification [19, 22]. The FDMM evaluates both data and metadata compliance with each FAIR data principle through one or more indicators [19]. Each indicator is associated with (often domain-dependent) impact levels described as essential, important, or useful [14]. Communities have developed standardised FAIRification templates and workflows for their respective research domains [2, 5, 23–26]. For example, Jacobsen et al. developed a generic FAIRification workflow that has been instructive for the FAIRification of electronically captured data on vascular anomalies and COVID-19 [16, 27–29]. To accommodate differences in contexts and scientific practices, Sinaci et al. proposed a FAIRification workflow which applies restrictions considerate of the technical, ethical and legal requirements specific to health research data [17]. This workflow has shown promise in improving health research management outcomes in terms of time and costs [4].

The German Center for Diabetes Research (known locally as Deutsches Zentrum für Diabetesforschung - DZD) is a federal government and states funded national association that brings together experts to conduct translational research across the full spectrum of diabetes and metabolism [30]. In 2021 the DZD established a minimum data set known as the DZD CORE DATA SET (CDS), which is geared toward research projects in the fields of diabetes and metabolism research [31]. The DZD CDS enables the harmonization of data items, labels, definitions, and documentation across all the DZD clinical studies, which further enables data comparability between related studies. The effort involved in conducting our recent retrospective FAIRification work informed the decision to employ structured FAIRification workflows in the FAIRification of the DZD CDS [12, 32]. The enhanced data sharing that has been facilitated by the FAIRification of the DZD CDS is expected to contribute to its increased uptake among relevant stakeholders within the DZD and the wider diabetes research community [12]. The objective of this work is to explore the retrospective implementation of a generic [16] and a domain-specific [17] FAIRification workflow using the DZD CDS as a case study. Stakeholders considering the implementation of FAIRification workflows may use the experiences and results obtained from this work to inform the development of FAIRification standard operating procedures (SOPs) for their research domains.

## Methods

### Data set

The DZD CDS is a mandatory component for the design of all upcoming DZD clinical studies and it has been implemented in various DZD studies since its conception [33–35]. It also includes the common CDS that was established among the German Centers for Health Research (*Deutsche Zentren der Gesundheitsforschung*) [36]. It contains 147 data items that have been selected by a group of medical experts and categorized into eight modules. It also contains optional modules relevant for special studies as shown in the following Figure 1.

**Fig 1.**
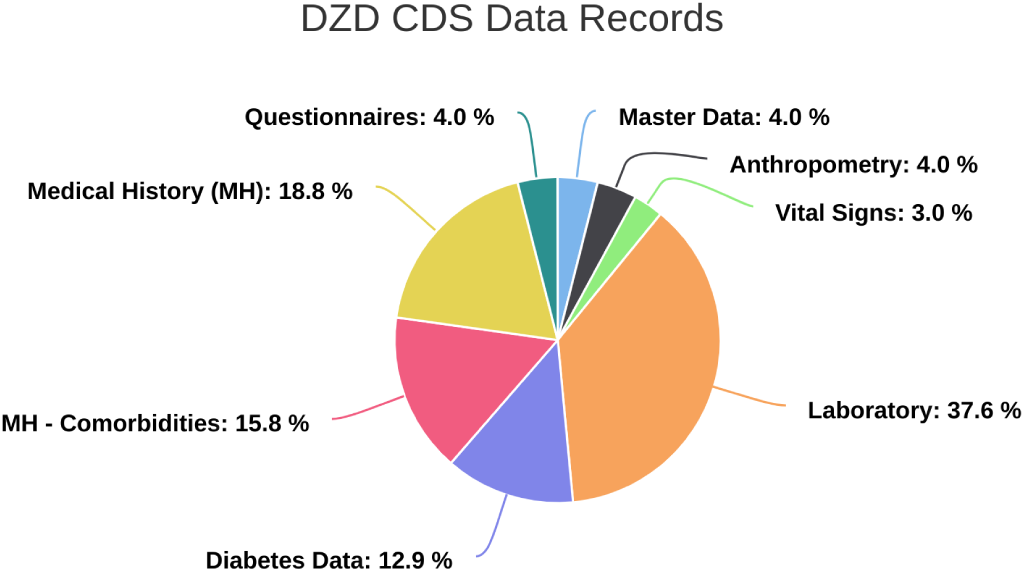
DZD CDS Data Records (Base Set). Included modules: master data (contains patient information), anthropometry (contains patients’ height, weight, waist & hip circumference, as well as the techniques used to obtain these measurements), vital signs (contains blood pressure and heart rate), laboratory (contains selected laboratory blood, plasma, serum and urine tests), diabetes data (contains date and type of diabetes diagnosis as well as treatment offered), medical history (contains a record of a patient’s health background and the occurrence of disease events), medical history-comorbidities (indicates the presence of co-existing diseases with reference to the diagnosis) and questionnaires (contains the Baecke Index for sports, leisure, work and the total score) [12].

### Analysis setup

A structured comparison was performed by retrospectively implementing the generic FAIRification workflow and the FAIRification workflow for health research in the context of the DZD CDS [16, 17]. The decision to employ the FDMM as an evaluation instrument for this work was arrived at after a series of tools were tried, tested and eliminated for various reasons such as poor interpretation of the FAIR data principles, poor FAIRness assessment methods, and poor user friendliness [37]. We also considered the success reported by other researchers who used this tool for their own FAIR assessments [14, 38–41]. The FDMM allowed us to discard the data indicators and instead focus on the metadata indicators (26 out of 41 indicators) as is necessary for the FAIR assessment of a CDS. Two data curators (ETI and AD) collaboratively assessed the DZD CDS FAIRness based on the publicly available guidance provided for this tool [19]. The curators assessed the DZD CDS FAIRness in three iterations for surety. In our assessment of the CDS we only used the FDMM metadata indicators shown in Table S1 (supporting information).

### Selection of FAIRification Workflows

The results of our previous scoping review of FAIRification workflows [5] led us to select the workflows developed by Jacobsen et al. and Sinaci et al. for the FAIRification of the DZD CDS [16, 17]. Both workflows have already been successfully implemented in various health research settings [4, 27, 28].

### Selection of a FAIR metadata repository

We included a structured selection process for a FAIR metadata repository in our FAIRification process prior to the implementation of the workflows. More specifically, we embarked on a search for a FAIR repository in which we would house the DZD CDS as part of the FAIRification. The Swiss National Science Foundation (SNSF) released a checklist for the selection of FAIR repositories [42].

Using the criteria from the SNSF checklist, we evaluated the Medical Data Models (MDM) Portal [43, 44] against the SNSF checklist for FAIR repositories. Table 1 shows the results of this evaluation.

**Table 1.** Adherence of the German Portal for Medical Data Models (MDM Portal) to the SNSF’s Criteria for FAIR Repositories [42].

| Repository Criteria | MDM Portal |
| --- | --- |
| Is the repository non-commercial? | The MDM Portal serves as an infrastructure for academic (non-commercial) medical research. |
| Are datasets (or ideally single files in a dataset) given globally unique identifiers (IDs) and persistent IDs? | The MDM Portal offers digital object identifiers (DOIs) as globally unique and persistent IDs which can be assigned after users' request. The MDM Portal also provides guidelines for its citation in publications, presentations and registration of studies in study registries such as Clinical-Trials.gov [45]. |
| Does the repository allow the upload of intrinsic and submitter-defined metadata? | Yes, basic intrinsic data has to be entered in specific mandatory fields. "Free text" fields are also available for the user to enter additional information. |
| Is it clear under which licence the data will be available, or can the user upload/choose a licence? | The user may upload the data models under one of four different Creative Commons Licenses including [46]:<br>• Creative Commons BY-NC 4.0<br>• Creative Commons BY-NC-SA 4.0<br>• Creative Commons BY-SA 4.0<br>• Creative Commons BY 4.0<br>The terms and conditions of these licenses have been stipulated in the portal for the submitter's reference. |
| Are the citation information and metadata always (even in the case of datasets with restricted access) publicly accessible? | Yes |
| Does the repository provide a submission form requesting intrinsic metadata in a specific format (to ensure machine readability)? | Yes, basic intrinsic metadata has to be entered in a structured way (online form) in required fields. Checks are performed on some fields to ensure that the proper format is used (for example, email address and date). |
| Is there a long term preservation plan for the archived data? | Yes |

Based on the results of this evaluation, we identified the MDM Portal as a FAIR repository employable in the FAIRification of the DZD CDS.

## Results

### Implementation of FAIRification Workflows

#### 1. Generic FAIRification workflow

The generic FAIRification workflow developed by Jacobsen et al. consists of 7 steps which are categorised into the pre-FAIRification phase, the FAIRification phase and the post-FAIRification phase [16].

##### Identifying the FAIRification objectives

We conducted a baseline FAIR assessment on the DZD CDS and then considered the results of this assessment (shown in Figure 2), along with the DZD CDS context, stakeholders’ priorities and the desired scientific value of the data to formulate the objectives of the DZD CDS FAIRification as follows:

- Findability: To improve the searchability and findability of the DZD CDS items for users and across future DZD CDS versions
- Accessibility: To maintain the DZD CDS items in a manner that allows for them to be accessed under well-defined access conditions
- Interoperability: To annotate the DZD CDS items with biomedical ontologies, data standards, terminologies and a structured format so as to facilitate interoperability and automatic extraction of relevant data items across different future DZD CDS versions
- Reusability: To represent the data in a concise manner that allows for reuse of the data collected across all the different future DZD CDS versions

**Fig 2.**
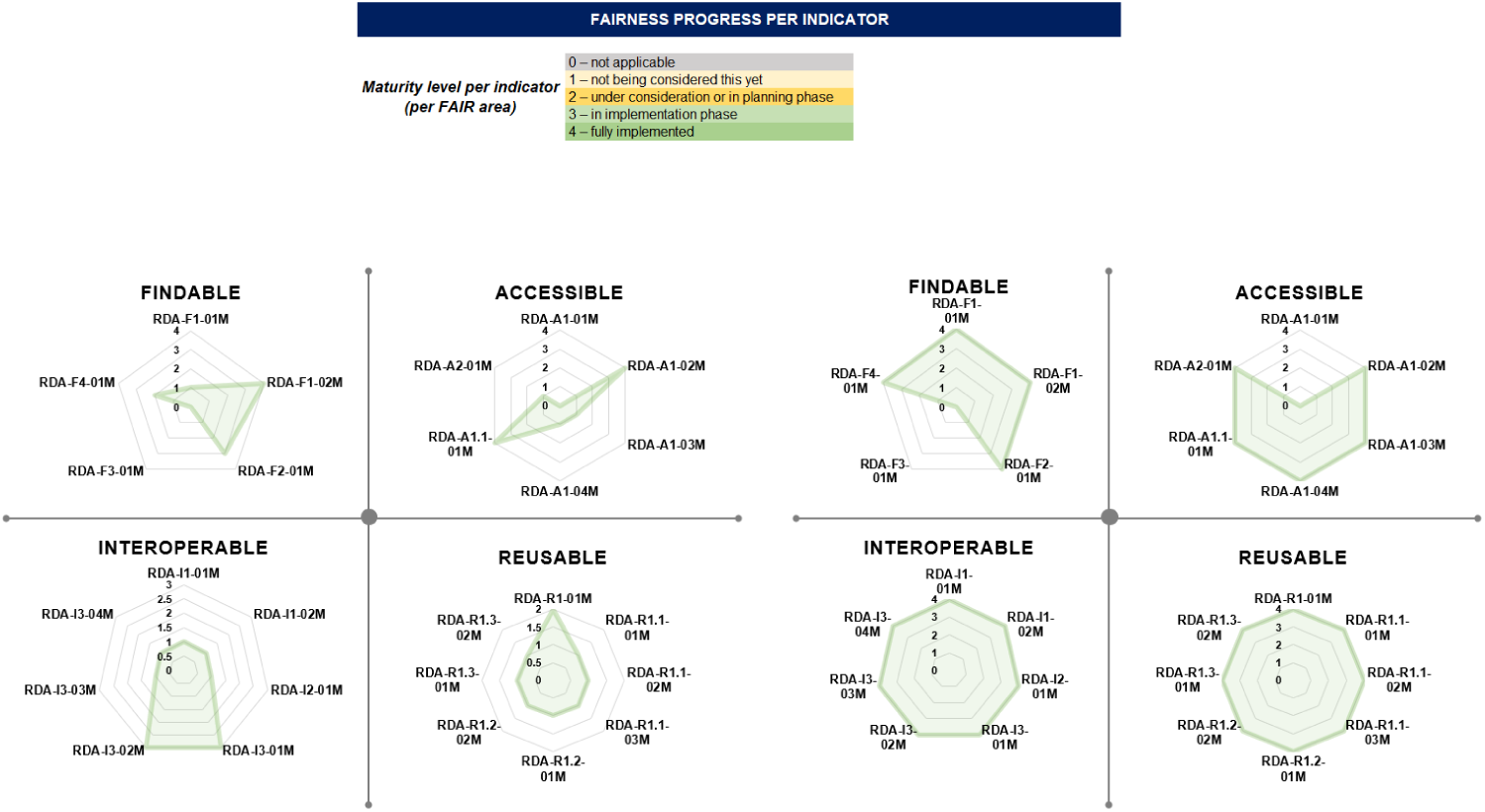
The diagram on the left illustrates the results of the baseline DZD CDS FAIR assessment while the diagram on the right illustrates the results of the final DZD CDS FAIR assessment. The indicator maturity levels are defined in the legend above the radar charts. This legend is used to indicate the FAIRness progress per indicator [12].

##### Analysis of the data and metadata

The data fields, types, and values were characterized. The data elements and data fields were extracted and the curated data was validated by the clinical data experts. We have identified missing metadata with regard to temporal and spatial factors as well as contact persons, keywords and information on the target group.

##### Defining the semantic data and metadata model

We used the version of the DZD CDS provided by the MDM Portal which offers a bottom-up standardization process that facilitates the semantic enrichment of all the data items with codes from LOINC, SNOMED CT and UMLS [44, 47, 48]. All previous versions of the DZD CDS are listed under https://medical-data-models.org/46011. **Making data and metadata linkable**: Contextual knowledge (persistent IDs and references to other data sets/publications) was added to the dataset in the form of meaningful links. The metadata and SOP were registered and hosted in Zenodo and the related Zenodo link was also added to the dataset [33, 49]. **Hosting FAIR data**: The DZD CDS has been hosted on the MDM Portal for purposes of making it a community resource available for human and machine consumption. Hosting the DZD CDS on the MDM Portal allows downloading and exporting the file in most common technical formats. Hosting the CDS on the MDM Portal also required us to assign it a licence that stipulates its reuse. In all DZD multicentred clinical trials, the direct identifying data (IDAT) are handled spatially and organisationally separated from the medical data (MDAT) to comply with legal, organisational and technical requirements regarding data protection [50]. For this reason the IDAT is not part of the CDS. This, along with the fact that the DZD CDS does not contain personal data, influenced the decision to choose an open machine-readable licence (the Creative Commons BY-NC-SA 4.0) [51]. Hosting the DZD CDS on the MDM Portal also led to data versioning and indexing.

##### Assessing FAIR data

Finally, we determined if the defined objectives have been achieved and conducted a final FAIR assessment. The final FAIR assessment results of the DZD CDS are shown in Figure 2.

Table S2 (supporting information) illustrates the implementation of the generic FAIRification workflow in the DZD CDS and the decisions made in adapting it to this context.

### FAIR assessments

We assessed the FAIRness of the DZD CDS before (left diagram) and after (right diagram) application of the FAIRification workflows using the FDMM. The results are shown in the following Figure 2.

We observe that all FAIR indicators recorded significant improvement after the implementation of the FAIRification workflows. The implementation of both workflows led us to achieve the same FAIRness score. The DZD CDS contains neither patient identification data nor sensitive data. For this reason, the RDA-A1-01M and RDA-F3-01M indicators (defined in Table S1, supporting information) were not applicable [14].

#### 2. Domain-specific FAIRification workflow

The FAIRification workflow specific to health research developed by Sinaci et al. consists of 10 steps [17].

##### Analysing the data and metadata

The implementation of this workflow began with raw (meta)data analysis similar to the one performed in the implementation of the generic FAIRification workflow.

##### Curating and validating the data

This step was performed as already indicated in the implementation of the generic FAIRification workflow.

##### De-identifying and pseudonymising data

We skipped the pseudonymization and de-identification step of this workflow because the DZD CDS does not contain any sensitive patient data.

##### Semantic modelling

This step was performed as already indicated in the step that calls for “definiting of the semantic data and metadata model” in the generic FAIRification workflow.

##### Making data and metadata linkable

The data was then enriched by adding contextual knowledge in the form of meaningful links as performed in the implementation of the generic workflow.

##### Attributing a licence, data versioning and indexing

These steps were already performed in the implementation of the generic workflow when the DZD CDS was registered in the MDM Portal. The same was done for the metadata and SOP in Zenodo where it was registered as shown in the implementation of the generic workflow [49].

##### Aggregating the metadata

The metadata was then aggregated, leading to the provision of the DZD CDS readme file and provenance information that contains the data origin, citations for reused data, description of the data collection, data processing history and version history.

##### Publishing

The DZD CDS was then published in the MDM Portal for consumption by the audience.

Table S3 (supporting information) illustrates the implementation of the health research FAIRification workflow in the DZD CDS and the decisions made in adapting it to this context.

### Similarities in Workflows

The following Figure 3 illustrates the steps taken in both FAIRification workflows and the principles achieved by implementing each step. The steps with similar colours are representative of the steps we found to be similar in our implementation. The bottom sketch shows our implementation of these workflows in the FAIRification of the DZD CDS.

**Fig 3.**
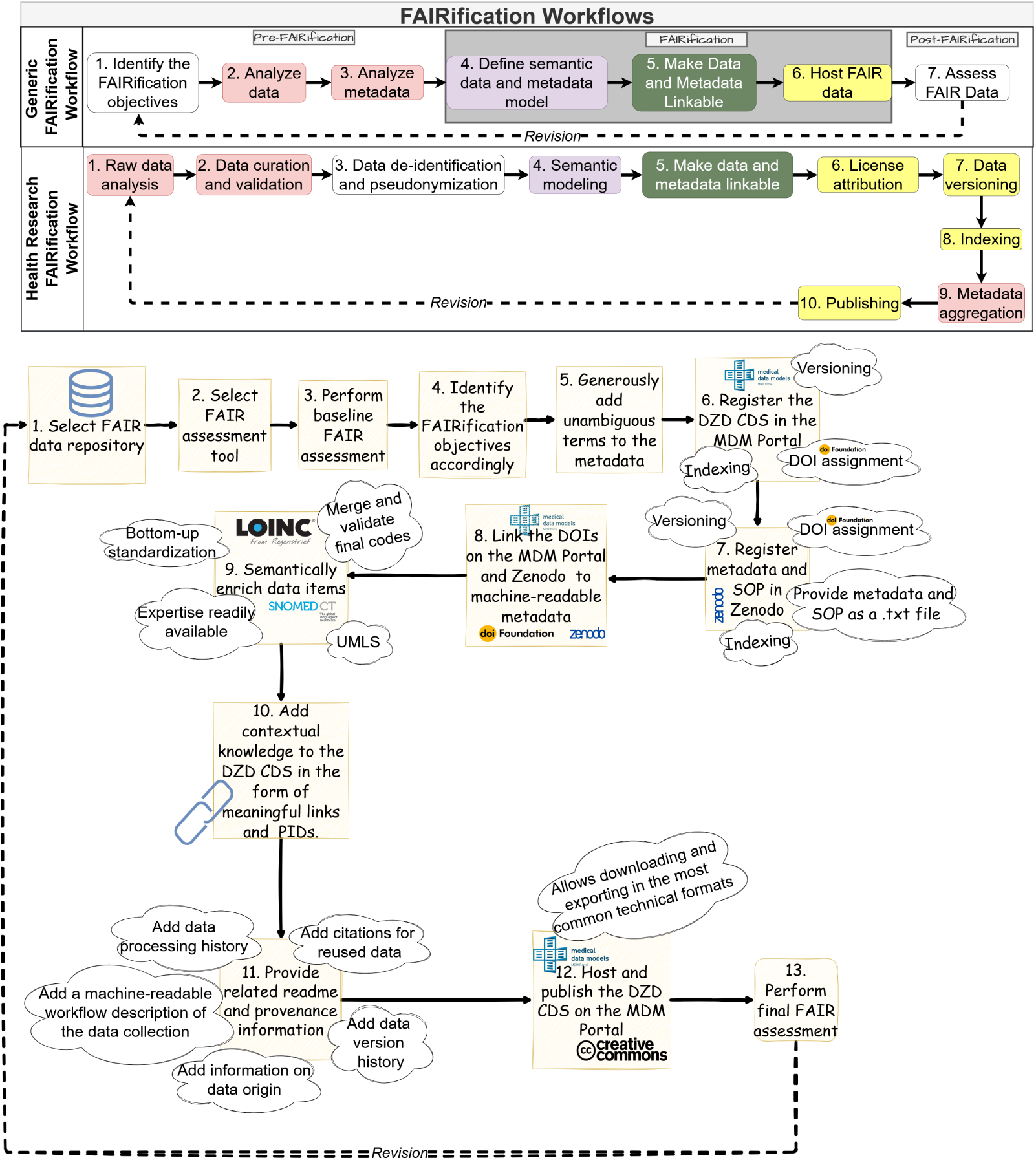
Comparison of generic and domain-specific worklows: The upper lane shows the generic workflow (Jacobsen et al.) and the lower lane shows the steps specific to the health research workflow (Sinaci et al.) [16, 17]. The sketch at the bottom shows our implementation of these workflows in the FAIRification of the DZD CDS.

Notably, information about licensing options are more distinct in the health research workflow than in the generic FAIRification workflow. Although the generic FAIRification workflow does not explicitly indicate licensing as a FAIRifcation step, we still licensed the CDS in our implementation of this workflow because hosting it on the MDM Portal required us to do so. Hosting the DZD CDS on the MDM Portal also simultaneously led to its versioning and indexing.

### Minimum Requirements for Successful FAIRification of core data sets in health research

This work has shown that regardless of the selected FAIRification workflow, there are some bare minimums that should be present for successful metadata FAIRification of core data sets in health research. They include:

1. A thorough understanding of the processes by which the metadata is defined, adapted, and expanded: The data analysis step helps to understand the project capabilities and resources. This step is also useful in developing an understanding the processes by which the dataset is defined, adapted, and expanded. It also helps to identify the data characteristics that should be improved based on the defined FAIRification goal. The capabilities that a FAIR data management environment should exhibit to enable and support the realization of a FAIR dataset can also be determined, including data access, data hosting, ontology services and data sharing. Finally, the findings of the raw data analysis play a critical role in determining the FAIR assessment tool and requirements needed to achieve the desired FAIRification outcome.
2. Collaborative development of standards: A cultural shift is required to enable the implementation and co-development of standards for metadata, centrally managed and organized [22, 52]. This cultural change needs to be broadly accepted in order to implement continuous FAIR RDM throughout the data lifecycle [26, 52].
3. Collaborative data stewardship: A shift away from the culture of individualised data ownership towards one of collaborative data stewardship is necessary for efficient data sharing that facilitates impactful research [53, 54]. Consultations and training for data publication pipelines should be offered to ensure sustainably operated FAIR infrastructures.
4. An investment of resources: A significant investment of time, money and domain knowledge is necessary to implement the described FAIRification steps [22, 55]. Customised incentives are important to encourage stakeholders to engage in data sharing beyond their moral obligations, especially if tangible motivators for this remain limited [54].

## Discussion

The comprehensive approach including initial FAIR assessments, FAIRification activities, post-FAIRification adjustments, and being intentional about implementing FAIR-enabling infrastructure can significantly improve the FAIRness of health data sets, as demonstrated in this work using the example of the DZD CDS. The application and comparison of two common FAIRification workflows underscores both the adaptability and domain-specific nuances of such processes. The domain-agnosticism of the generic workflow provides a flexible framework applicable to diverse datasets. Conversely, the health-specific workflow addresses the vital additions that are necessary to safeguard the integrity of health research data.

The inclusion of metadata aggregation and systematic data versioning in the health-specific workflow emphasizes the importance of robust data management practices to address the complexities of health research data. Domain-specific requirements shape FAIRification, balancing technical interoperability with adherence to contextual and legal constraints. Thereby, tailored health FAIRification workflows enhance the applicability of the FAIR principles and showcase how FAIRification efforts can be extended in other contexts, ensuring flexibility and precision.

The implementation of structured FAIRification workflows significantly improved the FAIRness of the DZD CDS. Previous FAIRification efforts, guided by experience rather than structured workflows, required extended timelines due to multiple consultations and iterative revisions [12, 56]. Contrarily, the two workflows applied in this work were straightforward to implement, with the FDMM proving instrumental. The FDMM facilitated both binary and scaled approaches to FAIR assessment, enabling objective evaluations with minimal reliance on personal judgement while allowing for progress measurement towards FAIRer scores. Baseline and final FAIR assessments, though not explicitly included in the FAIRificaton workflow for health research, were invaluable in evaluating improvements and demonstrating the impact of FAIRification efforts [13, 41]. Interestingly, the implementation of both workflows led us to achieve the same FAIRness rating. We therefore deduce that the adoptions made in the FAIRification workflow for health research data are useful to improve efficiency but do not necessarily lead to higher FAIRness scores when applied to core data sets.

Both workflows required similar resources for implementation but adapting them to specific objectives and contexts may improve cost and time efficiency, particularly when applied to real-world medical data. De-identification and pseudonymization are measures implemented to preserve the data privacy rights of the subjects and is performed based on the purpose for which the dataset has been developed [57, 58]. Unlike the generic FAIRification workflow, the FAIRification workflow for health research includes pseudonymization and de-identification as a distinct FAIRification step. This indicates workflow’s consideration of the heightened sensitivity of health research data, especially in the wake of the implementation of the General Data Protection Regulation in the European Union [59, 60]. Skipping this step in the implementation of the FAIRification workflow for health research did not adversely affect the final results of the final FAIR assessment. This may indicate that de-identification and pseudonymization of health research data does not directly contribute to data FAIRness. Pseudonymization and de-identification processes have been described as slow and cumbersome and would likely increase the effort required in the FAIRification of DZD CDS-related clinical research datasets that contain patient information [57, 61–64]. We were able to incorporate all the other metadata indicators without modifying them.

### Goal-Oriented and Flexible Implementation

While the health research FAIRification workflow does not explicitly include goal setting, defining the objectives has shown to be a critical component of FAIRification [6, 58]. In this context, collaboratively setting objectives helped to ensure that the objectives are inclusive of the perspectives of the various pertinent stakeholders and served as a means to justify the expenditure of resources in the FAIRification exercise. The business interest that is expected to evolve as the number of exports from the MDM Portal increase also served as a factor that encouraged collaboration among the stakeholders. There are also previous success stories that served as reference points [65–71]. The objectives also served as the basis for feedback on the effectiveness of individual FAIRification tasks that can be used to determine the overall success of the process and set a clear endpoint for this FAIRification iteration. Jacobsen et al. indicated that the generic FAIRification workflow steps need not follow a strict sequence [72]. This flexibility allowed us to start the DZD CDS FAIRifcation by selecting FAIR infrastracture, evaluating the baseline FAIRness and identifying the FAIRification objectives accordingly (step 1). We then proceeded to enrich the metadata as informed by our metadata analysis (steps 2 and 3) and then registered the DZD CDS in the MDM portal (step 6). The related metadata and SOP were registered in Zenodo. After this, we semantically annotated the data items since the MDM Portal provided codes and and we were able to discern these codes (Step 4). The semantic enrichment of DZD CDS raw data items with codes from LOINC, SNOMED-CT, and UMLS was conducted retrospectively, which presented a substantial clerical burden [48, 73]. Once this process was complete, subsequent workflow implementation (shown in Figure 3) required comparatively less time and effort.

### FAIR Infrastructure

Registering the DZD CDS in the MDM Portal alongside metadata and SOPs in Zenodo enabled simultaneous completion of multiple workflow steps as shown in Figure 3. This approach streamlined the process and demonstrated the importance of selecting FAIR infrastracture prior to the onset of a FAIRification journey. Jacobsen et al. have highlighted that the generic FAIRification workflow specifically targets the implementation of principles F1, F2, R1, R1.1, R1.2 and R1.3 [1, 72]. In our context these principles target the corresponding metadata sub principles as defined by the RDA (see Table S1, supporting information). Depositing the metadata and SOPs in Zenodo also led to the fulfillment of the corresponding F3 and F4 metadata principles (defined in Table S1, supporting information). Registering the DZD CDS in the MDM Portal led to the fulfillment of the “accessibility” metadata sub-principles. The DZD CDS can now be downloaded and exported in most common technical formats and provides UMLS codes for semantic enrichment, which further enables the implementation of an ontology matching service for querying FAIR data [12, 74]. Qualified references are available in the form of links to the comprehensive metadata and SOP in Zenodo which further fulfils the “interoperability” sub-principles. An interesting finding in the implementation of the generic workflow is that key FAIRification measures were already implemented while we were still in the pre-FAIRification phase. These include addition of rich explicit metadata, licensing, versioning, DOI assignment, as well as registration and indexing of the metadata and SOPs. This raises the question, “when does FAIRification actually begin?”

### Expected Impact of a FAIRer DZD CDS

FAIRification has increasingly become an important part of the DZD RDM priorities and this has necessiated FAIR data sharing of the DZD CDS. The FAIRer DZD CDS has been made available to the community on the MDM portal and supports data sharing among DZD sites [75]. FAIRification of the DZD CDS aligns with the DZD’s commitment to comprehensive data stewardship [12], enhanced data sharing, and data analysis across DZD sites [12, 56]. A machine-actionable framework to describe and structure the CDS has been established and the DZD CDS is more interoperable [48, 76]. The expected return on the investment of the efforts made to provide machine-actionable, heterogenous data is that there are now wider possibilities for data integration and larger-scale analyses. Expanding access via alternative authentication and authorisation procedures could further enhance utility for non-MDM portal users.

This work addresses only a single CDS. Therefore we cannot comment on any domain-specific challenges nor had any particular domain-specific requirements that influenced the FAIRification process. We expect, however, that such challenges may occur in the context of real-world data related to the DZD CDS.

### Is the FAIRification Journey of Medical Research Data Worthwhile?

FAIRification of medical research data is essential for evidence-informed decisions and has proven beneficial for datasets such as the DZD CDS, which has garnered significant interest (725 views and 379 downloads as of November 2024) [5, 77, 78]. Enhanced reusability increases the value of the CDS, provides a basis for profitable reuse in other contexts, and may eliminate the need for a new data collection process. The improved reusability of the data set over an extended period of time may further result in the development of new therapeutic regimens by the secondary data user and increased business value for the DZD.

Interoperability, supported by standardized technical formats and metadata, supports integration with heterogeneous datasets, broadening research opportunities, and simplifying cross-evaluation [79]. Since all current DZD clinical studies launched in 2021 and later use the same CDS, the data pool is enlarged and the possibility of cross-evaluation is simplified [12]. Novel research based on integrated and analyzed heterogenous data is anticipated once the FAIRified DZD CDS is connected to routine data from data integration centers [80, 81]. Taken together, these efforts improve discoverability and readiness for artificial intelligence, contributing to greater visibility and impact of the DZD CDS.

The key resources required for this iteration of the DZD CDS FAIRification included funding, expertise, and incentives. The DZD CDS FAIRification required numerous consultations and meetings between data owners and FAIR experts to make key decisions, which contributed to a significant time investment. The quantified investment amount that should be made in running FAIRification cycles in a manner that keeps it beneficial to stakeholders remains an open question. We recommend that this discussion should include the stakeholders of other clinical core data sets such as the one developed by the German Medical Informatics Initiative and the French CDS for geriatric oncology studies [32, 82, 83].

## Conclusion

In this work, we compared the implementation of two FAIRification workflows for the core data set applied to German diabetes studies. We also identified minimums that will help to reduce efforts and costs for FAIRification, when applied. We recommend that more FAIRification workflows that take into account the nature of domain-specific data should be developed for other scientific domains that have embraced FAIRification as a necessary journey.

Retrospective data FAIRification is a cumbersome task regardless of the FAIRification workflow implemented. For this reason, we resonate with current recommendations that encourage scientists and data owners to design their scientific projects in a way that takes into account the FAIRification of prospective data right from the infancy stage and continuously improves FAIRness throughout the lifecycle of the project [22, 84]. One aspect of these preparatory steps is the thoughtful collection of appropriate FAIRification tools (infrastructure) as early as possible.

However, this is a multi-stakeholder engagement and different stakeholders may have different preferences with regards to the FAIRification process. For example, the funders may deem it more cost-effective to FAIRify data all in one cycle as opposed to gradually and iteratively while other participant stakeholders may have different preferences on the order in which the FAIRification steps should be implemented. The implementation of both FAIRification workflows in the DZD CDS would not have been possible without the tremendous involvement of the data owners, data stewards and the pertinent stakeholders. We therefore recognise the importance of harmonizing the stakeholders’ perspectives and expectations.

Templates for FAIR data management plans (DMPs) continue to be developed as funders and policy makers continue to require DMPs to be prospectively FAIR inclusive [5, 25, 42, 85]. It remains to be seen how these prospective FAIR DMPs can be integrated into retrospective FAIR workflows. It may also be necessary to develop FAIRification workflows for prospective implementation specific to the health research data domain. It also remains to be explored what the resultant differences would be to the FAIRification workflows if the respective steps were implemented prospectively.

Quite a lot of work has already been done to automate the semantic enrichment of health data [86–89]. It would be interesting to further research how automated semantic enrichment can be incorporated into FAIR workflows, how many more of the steps in the FAIR workflows can be automated, and what would be the consequent changes, if any, to the workflows once the steps are automated. In 2019 the European Commission estimated the cost of not having FAIR research data in the European research economy at € 10.2 billion [90]. Another interesting area of research would be to determine how much time and resources has been saved by FAIRifying the DZD CDS. It remains to be seen which steps will be iterated or eliminated in subsequent FAIRification cycles as the priorities of this FAIRification journey evolve and new insights are obtained.

## Data Availability

An archived record of the previous version of the DZD CDS before FAIRification is
retrievable in Zenodo at: https://doi.org/10.5281/zenodo.12526690.
The FAIRified version of the DZD CDS has been deposited in the MDM Portal and is
retrievable at: https://medical-data-models.org/46011.
The related metadata and SOPs have been deposited into Zenodo and is retrievable
at: https://zenodo.org/record/7360000.

https://medical-data-models.org/46011

https://zenodo.org/record/7360000

## Supporting information

An archived record of the previous version of the DZD CDS before FAIRification is retrievable in Zenodo at: https://doi.org/10.5281/zenodo.12526690 [91].

The FAIRified version of the DZD CDS has been deposited in the MDM Portal and is retrievable at: https://medical-data-models.org/46011 [31].

The related metadata and SOPs have been deposited into Zenodo and is retrievable at: https://zenodo.org/record/7360000 [33].

**S1 Table. FDMM Metadata Indicators**

Table S1 illustrates the RDA FDMM metadata indicators that we employed in the FAIRness assessment of the DZD CDS [14].

**S2 Table. Implementation of the generic FAIRification workflow**

Table S2 illustrates the implementation of the generic FAIRification workflow in the DZD CDS and the decisions made to adapt it to this context [16].

**S3 Table. Implementation of the health research FAIRification workflow**

Table S3 illustrates the implementation of the health research FAIRification workflow in the DZD CDS and the decisions made in adapting it to this context [17].

## Acknowledgements

This work was partially funded by the NFDI4Health – Nationale Forschungsdateninfrastruktur für personenbezogene Gesundheitsdaten (DFG-funded project 442326535) and the Deutsches Zentrum für Diabetesforschung (German Center for Diabetes Research).

