## Supplementary Information for "Lessons learnt from implementing FAIRification workflows in diabetes research in Germany"

The following Table S1 shows the FDMM metadata indicators used in the FAIR assessment of the DZD CDS.

| Indicator ID | Indicator | Priority |
|---|---|---|
| RDA-F1-01M | Metadata is identified by a persistent identifier | Essential |
| RDA-F1-02M | Metadata is identified by a globally unique identifier | Essential |
| RDA-F2-01M | Rich metadata is provided to allow discovery | Essential |
| RDA-F3-01M | Metadata includes the identifier for the data | Essential |
| RDA-F4-01M | Metadata is offered in such a way that it can be harvested and indexed | Essential |
| RDA-A1-01M | Metadata contains information to enable the user to get access to the data | Important |
| RDA-A1-02M | Metadata can be accessed manually (i.e. with human intervention) | Essential |
| RDA-A1-03M | Metadata identifier resolves to a metadata record | Essential |
| RDA-A1-04M | Metadata is accessed through standardized protocol | Essential |
| RDA-A1.1-01M | Metadata is accessible through a free access protocol | Essential |
| RDA-A2-01M | Metadata is guaranteed to remain available after data is no longer available | Essential |
| RDA-I1-01M | Metadata uses knowledge representation expressed in standardized format | Important |
| RDA-I1-02M | Metadata uses machine-understandable knowledge representation | Important |
| RDA-I2-01M | Metadata uses FAIR-compliant vocabularies | Important |
| RDA-I3-01M | Metadata includes references to other metadata | Important |
| RDA-I3-02M | Metadata includes references to other data | Useful |
| RDA-I3-03M | Metadata includes qualified references to other metadata | Important |
| RDA-I3-04M | Metadata include qualified references to other data | Useful |
| RDA-R1-01M | Plurality of accurate and relevant attributes are provided to allow reuse | Essential |
| RDA-R1.1-01M | Metadata includes information about the licence under which the data can be reused | Essential |
| RDA-R1.1-02M | Metadata refers to a standard reuse licence | Important |
| RDA-R1.1-03M | Metadata refers to a machine-understandable reuse licence | Important |
| RDA-R1.2-01M | Metadata includes provenance information according to community-specific standards | Important |
| RDA-R1.2-02M | Metadata includes provenance information according to a cross-community language | Useful |
| RDA-R1.3-01M | Metadata complies with a community standard | Essential |
| RDA-R1.3-02M | Metadata is expressed in compliance with a machine-understandable community standard | Essential |

**Table S1.** **FDMM Metrics Indicators**

The following Table S2 illustrates the implementation of the generic FAIRification workflow in the DZD CDS and the decisions made in adapting it to this context.

| Step | Implementation |
|---|---|
| 1. Identify the FAIRification objectives | We considered the results of the baseline FAIR assessment, the DZD context and priorities, as well as the intended scientific value of the data, to formulate the objectives of the DZD CDS FAIRification as follows:<br>• Findability: To enhance the searchability and discoverability of DZD CDS items for users and across future versions.<br>• Accessibility: To ensure that DZD CDS items can be accessed under well-defined and transparent access conditions.<br>• Interoperability: To annotate DZD CDS items using biomedical ontologies, data standards, and terminologies, and to structure them in a way that facilitates interoperability and the automated extraction of relevant items across future DZD CDS versions.<br>• Reusability: To represent the data in a concise, standardized format that supports reuse across all future versions of the DZD CDS.<br>An improved FAIRification score serves as the primary benchmark for the success of this work. |
| 2. Analyze Data | The goal of this step is to analyze the data to prepare them for subsequent FAIRification and reinforce step 1 of this workflow. The DZD CDS data types include dates, integers, text (string), floats and booleans. In line with the goal of this step, we applied FDMM to assess the baseline FAIRness of the DZD CDS. We only considered the FDMM metadata indicators in the FAIR assessment of the DZD CDS. This adaptation was necessary to fit the FAIR assessment to the context of a CDS. The results of this assessment are shown in Figure 2. |

*Continued on next page*

| Table S2 – continued from previous page | |
|---|---|
| **Step** | **Implementation** |
| 3. Analyze Metadata | The availability of the necessary metadata as per stakeholder requirements was investigated. Prior to this FAIRification work, the DZD was accessible via a URL constructed using the Hypertext Transfer Protocol. The DZD CDS-related metadata consisted only of the title and names of individuals who had contributed to the dataset to some extent. However, the metadata lacked persistent unique identifiers and did not include a plurality of accurate and relevant attributes. Moreover, it was neither registered nor indexed in a searchable resource, and it was not maintained in a machine-exchangeable format. The metadata was therefore insufficient. The following were identified:<br>• The structure and relationships within and among the data element concepts<br>• The instructions for correct data collection for the following data items: height, weight, measurement of waist and hip circumference, laboratory examination data, equipment and materials required for the collection of the DZD CDS data<br>• The recommended workflow for the collection of the data contained in the DZD CDS (including the respondent interview, physical examination and medical laboratory examinations and the documentation protocol of the collected data)<br>• The implemented clinical data models, coding schemes, terminology systems.<br>As a result of this analysis, unambiguous terms were generously added to the metadata. Detailed work instructions and metadata for all data items were stored in the MDM Portal, in accordance with the metadata schema for clinical research provided there. The metadata was enriched with contextual, spatial, and temporal coverage information; descriptions of the target audience; descriptive keywords; a license stipulating data access and reuse conditions; references to related datasets/resources; file formats used in the dataset; administrative metadata; and descriptive metadata for all data elements and concept properties.<br>The DZD CDS was registered and indexed in the MDM Portal under ID 46011, and a DOI was assigned [1, 2], enhancing its discoverability as a digital resource. The dataset was subsequently versioned and tagged with keywords, human-readable descriptions, and data types for each data element.<br>Additional metadata and a standard operating procedure (SOP) were registered in Zenodo, where they were versioned and assigned a DOI [3]. The metadata was also submitted to DataCite servers during DOI registration, where it was indexed [4].<br>Prior to the FAIRification process, the metadata and SOP were only available in PDF format on the DZD website. This effort resulted in their availability as machine-readable '.txt' files. The DOIs assigned in the MDM Portal and Zenodo were linked to machine-readable metadata, allowing identifiers to remain persistent even when semantic information changes [5, 6]. The inclusion of enriched metadata in this way significantly contributes to the increased visibility of the DZD CDS. |

*Continued on next page*

| Table S2 – continued from previous page | |
|---|---|
| **Step** | **Implementation** |
| 4a & 4b. Define Semantic Data and Metadata Model | The MDM Portal has a structure and process on how to assign codes known as bottom-up standardization which facilitated semantic enrichment of the data items with codes from LOINC and UMLS [7, 8]. SNOMED CT codes were also added [9]. This effort enables exchange between computer systems and ensures that the DZD CDS complies with community standardization efforts and is representative of a consensus view within the diabetes research community [9, 10]. All modules and items were successfully annotated. This led to the establishment of a well-defined framework to describe and structure the CDS in order to ensure its findability and interoperability. The codes from the MDM Portal were merged into a final set of codes and validated by the experts. In doing so the DZD CDS has been provided in a machine-readable format using an established and accessible format. This step allows for heterogeneous machines to retrieve, exchange, and aggregate data in one format. It also allows for the DZD CDS to be searchable and compatible data sources to be combinable in a (semi)automatic way. |
| 5a & 5b. Make Data & Metadata Linkable | This step requires that a description of the data and metadata is available in a representation framework that is globally understood by machines and that the semantic model is associated with the data and metadata so that it is available for unforeseen future applications and scalable interoperability across all types of data [11]. Contextual knowledge was added to the dataset in the form of meaningful links including PIDs. This step promotes interoperability and reuse, facilitating the integration of the data with other types of data and systems. This serves to transform the data to a FAIR representation. The metadata schema for clinical research provided in the MDM Portal was used. Related readme and provenance information was provided as follows:<br><br>• Origin of data, citations for reused data: Where available, it is found under "Description" at the MDM Portal entry. i.e. for the "Baecke Index: leisure index" [12]. All questions of the German version of the Baecke Score are part of the minimal dataset [13].<br><br>• Workflow description for collecting data (machine readable): The very detailed standard operating procedure has been published in Zenodo in PDF.<br><br>• The processing history of data and the version history of data: The first version (1.1.0) was published in 2021 as an excel sheet on the DZD website and is now hosted in Zenodo [14]. The 126 data items contained in the DZD CDS have since been harmonized between all DZD study centers to define a consistent data set within the DZD clinical studies. |

*Continued on next page*

| Table S2 – continued from previous page | |
|---|---|
| **Step** | **Implementation** |
| 6. Host FAIR Data | The DZD CDS has been hosted on the MDM Portal for purposes of making it a community resource available for human and machine consumption (`https://medical-data-models.org/46011`). The terms and conditions for reuse of this CDS were stipulated under an assigned license. In all DZD multicentred clinical trials, the direct identifying data (IDAT) are handled spatially and organisationally separated from the medical data (MDAT) to comply with legal, organisational and technical requirements regarding data protection. For this reason the IDAT is not part of the CDS. Record linkage is handled via a pseudonymization service offered by a trusted agency. This, along with the fact that the DZD CDS does not contain personal data, influenced the decision to choose an open machine-readable licence (the Creative Commons BY-NC-SA 4.0) [15].<br>Publishing the DZD CDS on the MDM Portal allows downloading and exporting the file in most common technical formats such as ODM, PDF, CDA, CSV, FHIR Questionnaire (JSON), FHIR Questionnaire (RDF), FHIR (ValueSet), FHIR Questionnaire (XML), NUM-Compass, MACRO-XML, Open Data Kit, OpenClinica, REDCap, ResearchKit, ResearchKit Swift, SQL, SPSS, ADL, R, XLSX etc. |
| 7. Assess FAIR Data | Implementation of this step included determining if the defined objectives have been achieved and a final FAIR assessment. The final FAIR assessment results of the DZD CDS are shown in Figure 3. |

Table S2. **Implementation of the generic FAIRification workflow in the DZD CDS**

The following Table S3 illustrates the implementation of the health research FAIRification workflow in the DZD CDS and the decisions made in adapting it to this context [16].

**Table S3: FAIRification Workflow for Health Research**

| **Step** | **Implementation** |
|---|---|
| 1. Raw Data Analysis | The result attained from conducting this step is similar to the result attained from conducting step 2 of the already demonstrated generic FAIRification workflow. |
| 2. Data Curation and Validation | The data fields, types, and values were characterized. The data elements and data fields were extracted and the curated data was validated by the clinical data experts. |
| 3. Data De-identification and Pseudonymization | In aligning this workflow to the nature of the DZD CDS, we skipped the step of data de-identification and pseudonymization because this dataset does not contain any information which would comprise the data subjects' rights regarding privacy issues. |
| 4. Semantic Modeling | The result attained from conducting this step is similar to the result attained from conducting step 4 of the already demonstrated generic FAIRification workflow. |
| 5. Make Data Linkable | The result attained from conducting this step is similar to the result attained from conducting step 5 of the already demonstrated generic FAIRification workflow. |
| 6. License Attribution | As shown in step 6 of the already demonstrated generic workflow, registering the DZD CDS led to the assignment of a license that stipulates reuse [2]. |
| 7. Data Versioning | As shown in step 3 of the already demonstrated generic workflow, registering the DZD CDS led to the dataset being versioned and tagged with keywords. Additional metadata and standard operating procedure (SOP) were registered in Zenodo where it was versioned [3]. |
| 8. Indexing | As shown in step 3 of the already demonstrated generic FAIRification workflow, the DZD CDS was indexed during its registration in the MDM Portal [2]. |
| 9. Metadata Aggregation | As shown in step 5 of the already demonstrated generic FAIRification workflow, this step led to the provision of the DZD CDS provenance and readme file. |

Continued on next page

Table S3 – continued from previous page

| Step | Implementation |
| --- | --- |
| 10. Publishing | The result attained from conducting this step is similar to the result attained from conducting step 6 of the already demonstrated generic FAIRification workflow. |

**Table S3. Implementation of the health research FAIRification workflow in the DZD CDS**
